# Zika epidemic in Colombia and environmental and sociodemographic contributors: an application of a space-time Markov switching model

**DOI:** 10.1101/2023.07.01.23292093

**Authors:** Laís Picinini Freitas, Dirk Douwes-Schultz, Alexandra M. Schmidt, Brayan Ávila Monsalve, Jorge Emilio Salazar Flórez, César García-Balaguera, Berta N. Restrepo, Gloria I. Jaramillo-Ramirez, Mabel Carabali, Kate Zinszer

## Abstract

Zika, a viral disease transmitted to humans by the bite of infected *Aedes* mosquitoes, emerged in the Americas in 2015, causing large-scale epidemics. Colombia alone reported 72,031 Zika cases between 31/May/2015 and 01/October/2016. We used national surveillance data from 1,121 municipalities over 70 epidemiological weeks to identify sociodemographic and environmental factors associated with Zika’s emergence, re-emergence, persistence, and transmission intensity in Colombia. We fitted a zero-state Markov-switching spatio-temporal model under the Bayesian framework, assuming Zika switched between periods of presence and absence according to spatially and temporally varying probabilities of emergence/re-emergence (from absence to presence) and persistence (from presence to presence). These probabilities were assumed to follow a series of mixed multiple logistic regressions. When Zika was present, assuming that the cases follow a negative binomial distribution, we estimated the transmission intensity rate. Our results indicate that Zika emerged/re-emerged sooner and that transmission was more intense in municipalities which were more densely populated, with lower altitude and/or less vegetation cover. Higher weekly temperatures and less weekly-accumulated rain were also associated with Zika emergence. Zika cases persisted for longer in more densely populated areas and with a higher number of cases reported in the previous week. Overall, population density, elevation, and temperature were identified as the main contributors of the first Zika epidemic in Colombia. The estimated probability of Zika presence increased weeks before case reporting, suggesting undetected circulation in the early stages. These results offer insights into priority areas for public health interventions against emerging and re-emerging *Aedes*-borne diseases.

## INTRODUCTION

Zika virus emerged in several tropical countries and territories, causing large epidemics between 2014 and 2016. As for dengue and chikungunya viruses, Zika is transmitted to humans by the bite of infected *Aedes* mosquitoes, mainly *Aedes aegypti*. These diurnal mosquitoes are well adapted to urban settings, live in intradomicile and peridomicile spaces, reproduce in small collections of fresh water, and are climate-sensitive. At the population-level, characteristics that vary in space (e.g. elevation) and in space and time (e.g. temperature) impact *Aedes aegypti*’s presence, density, activity, and competence to transmit viruses. Hence, they may also play a role in the spatio-temporal distribution of the diseases that are transmitted by *Aedes aegypti*, such as Zika.

Warmer temperatures were previously associated with an increased risk of *Aedes*-borne diseases ^1–5^. Up to 35^°^C, the increase in temperature increases the speed of *Aedes aegypti*’s life cycle (resulting in mosquito population growth) and the biting rates, while also decreases the extrinsic incubation period (which is the time taken between the mosquito ingesting the virus and becoming infectious) ^6,7^. By contrast, Zika transmission and vector competence is drastically reduced below 20^°^C ^7,8^. Humidity has been associated with the *Aedes aegypti* activity, survival and reproductive activity ^9,10^. Rainfall can fill small containers, such as any recipient from uncollected waste with fresh water, forming ideal breeding sites for the *Aedes* mosquitoes to lay their eggs and live during the immature stages ^9^. For that reason, improper waste management also plays an important role in the risk of diseases transmitted by *Aedes aegypti*, together with other factors that can increase the risk of exposure (e.g., housing materials, household crowding, access to piped water), particularly in areas with lower socioeconomic conditions ^1,11–13^. Finally, higher human population densities increase the chances of the mosquito-virus-human interaction.

Colombia has been endemic for dengue for decades, and the first Zika cases in the country were reported in August 2015^14,15^. Colombia has a very diverse geography, including islands, deserts, forests and mountain regions. Sociodemographic conditions also vary greatly within the territory. A robust National Surveillance System (*Sistema Nacional de Vigilancia en Salud Pública* - SIVIGILA) is implemented in the country, where all cases seeking care in health facilities with a probable or confirmed diagnosis of Zika must be reported ^16^. SIVIGILA data have been used to study the spatio-temporal patterns of *Aedes*-borne diseases (namely dengue, Zika, and chikungunya) in Colombia. However, most studies focused on a given municipality and/or department ^11,17–25^, and only a few considered the data for the whole country ^15,26–30^. Among these, the studies applying statistical models ^26–28^ used data at the department level.

One reason for the lack of nationwide spatio-temporal studies at the municipal level in Colombia is that across the territory, the distribution of *Aedes*-borne diseases is highly heterogeneous. Figure 1 displays examples of the different temporal patterns of Zika reported cases that can be observed across various geographically-distinctive Colombian municipalities. In some municipalities, there have never been reports of Zika cases. In others, the reported disease cases alternate between long periods of no cases followed by long periods of cases which are often interspersed with zero cases and occur at different magnitudes overtime. This complex type of data structure does not fit with most conventional statistical count models, such as Poisson or negative binomial regression models, due to the large number of zeroes ^31^. Also, while zero-inflated models can inflate the probability of observing a zero count ^32^, they would likely not be able to explain well the excessive numbers of consecutive zeroes ^33^.

**Figure 1:**
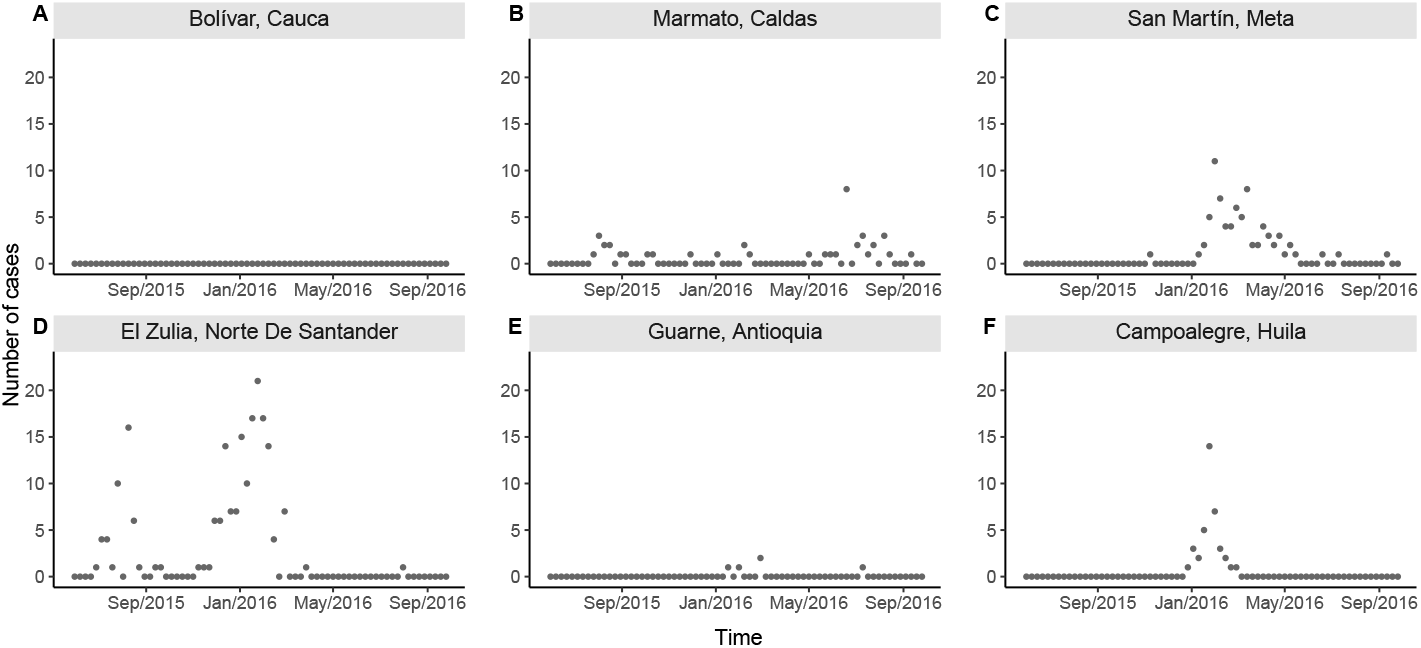
Examples of different temporal patterns of Zika reported cases counts by epidemiological week of first symptoms in selected municipalities of Colombia, epidemiological weeks 22/2015 to 39/2016: A) disease observed always absent in Bolívar (above 1800m of altitude and average temperature of 20^°^C); B) few cases reported intermittently in Marmato (above 1300m, 22^°^C); C) an observed emergence with very few cases followed by observed extinction and observed re-emergence with more cases being reported in San Martín (below 500m, 27^°^C); D) an early emergence followed by observed extinction and subsequent re-emergence in El Zulia (below 200m, 27^°^C); E) few and sporadic cases being reported in Guarne (above 2000m, 18^°^C); and F) an observed emergence followed by observed extinction in Campoalegre (around 500m, 24^°^C). Data source: Colombian National Public Health Surveillance System - *Sistema Nacional de Vigilancia en Salud Pública* (SIVIGILA).

In ^34^ they proposed a zero-state Markov switching model to address the challenging statistical task of modeling an excessive number of zeroes in spatio-temporal infectious disease counts. They assumed the disease switches between periods (i.e. *states*) of presence and absence in each area through a series of Markov chains. This usually means that when the disease is in a given state it is more likely to remain in that state. Therefore, their model can produce long strings of zeroes and long periods of positive counts, interspersed with zeroes, as is commonly observed in spatio-temporal counts of infectious disease cases. Also, unlike existing zero-inflated models, this approach allowed the distinction between the re-emergence (absence to presence) and persistence (presence to presence) of the disease which is epidemiologically justified in many instances ^34,35^. Our approach, explained in more detail below, extends ^34^ by adding an initial absence period to their model as we are modeling the initial introduction of Zika in Colombia. We used the model to explore how environmental and sociodemographic factors were associated with the emergence, re-emergence, persistence, and transmission intensity of Zika across the 1,121 municipalities of Colombia between 31/May/2015 and 01/October/2016. A diagram presenting our basic model structure is given in Figure 2. We assumed that Zika switched between periods of initial absence, presence, and subsequent absence in each of the municipalities according to spatially and temporally varying probabilities of Zika emergence, re-emergence, and persistence. These probabilities were assumed to follow a series of mixed multiple logistic regressions, allowing the estimation of associations with environmental and sociodemographic factors, such as temperature and population density, for example. When Zika was present in an area, the transmission intensity rate was modeled as a log-linear regression on environmental and sociodemographic factors. We recognize that zero reported cases can arise from either of the absence states, representing the true absence of the disease, or from the presence state, representing undetected Zika. Hence, an important aspect of our approach is that we can calculate the probability that a zero came from either of the *absence states* or from the *presence state* ^34^, and can therefore investigate where and when the disease was circulating undetected.

**Figure 2:**
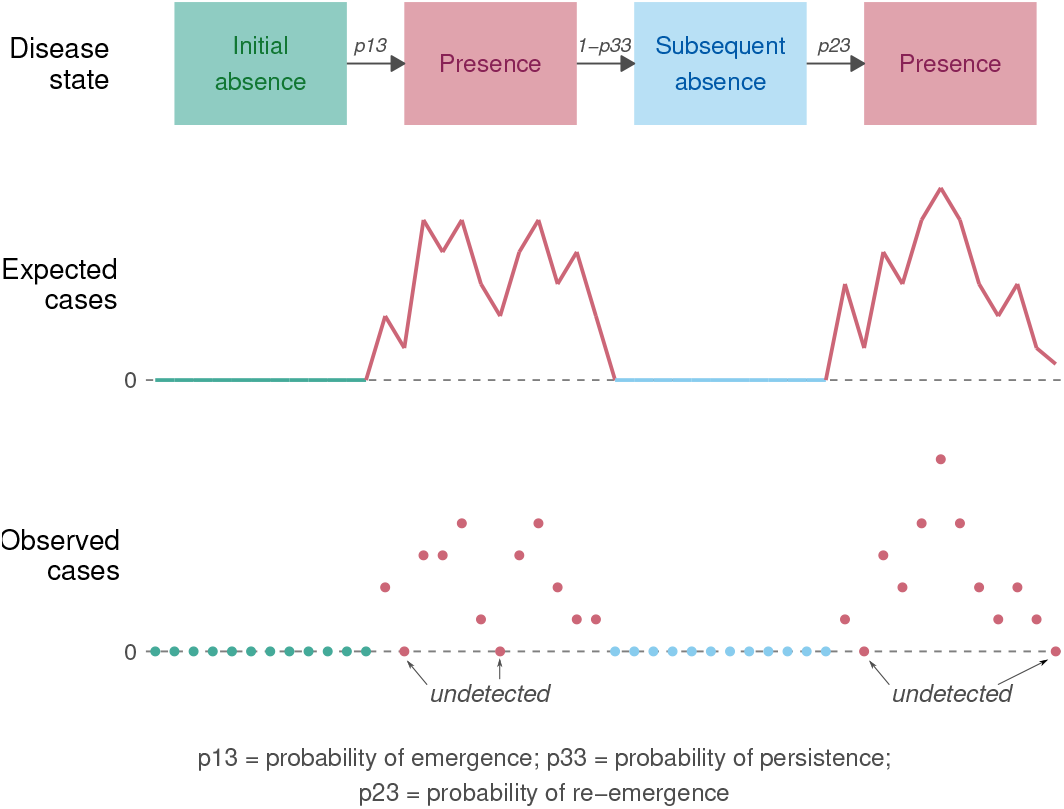
Diagram presenting the basic model structure considering three disease states: initial absence, presence, and subsequent absence.

## METHODS

In this ecological study, we analyzed the counts of Zika reported cases by municipality and week obtained from the Colombian National Public Health Surveillance System (*Sistema Nacional de Vigilancia en Salud Pública* - SIVIGILA).

### Study site

Colombia is located in South America and has 1,141,748 km^2^ and approximately 50.4 million inhabitants. The Colombian territory is divided into 1,121 municipalities that are grouped in 33 departments (Figures 3 and S1). The country borders with other five countries (Panama, Venezuela, Brazil, Peru and Ecuador), in addition to the Pacific Ocean to the west and the Caribbean Sea of the Atlantic Ocean to the north. Colombia’s geography is very diverse and can be classified into six main natural regions: the Andes mountains, the Pacific coast, the Caribbean coast, the Llanos (savanna), the Amazon rain forest, and the insular area. Colombian climate is considered tropical and varies across its natural regions.

**Figure 3:**
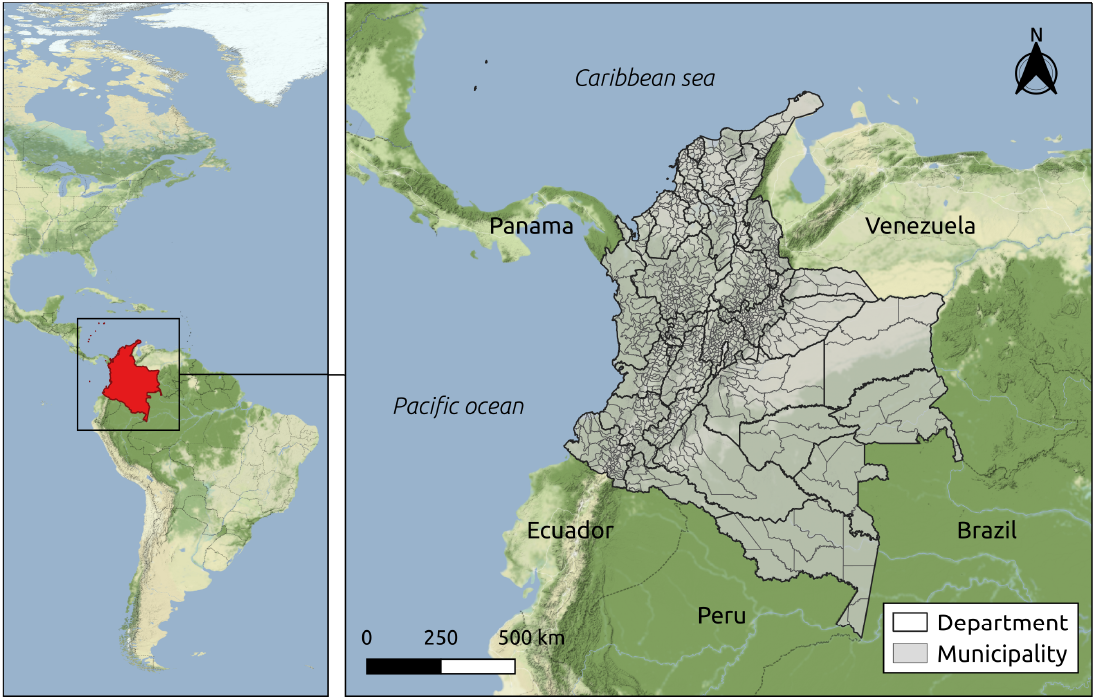
Localization of Colombia in the Americas and map of Colombia with the municipalities and geographical departments limits. (Sources: Colombian National Administrative Department of Statistics - DANE - geoportal. Background map tiles by Stamen Design, under CC BY 3.0. Data by OpenStreetMap, under ODbL.)

### Zika cases data

We obtained Zika reported cases data in all Colombia from the SIVIGILA portal ^36^. We aggregated the cases by the epidemiological week of symptoms onset and municipality of residence. The resulting cases dataset comprises the counts of probable and confirmed Zika cases over 70 epidemiological weeks and 1,121 Colombian municipalities during the period of the first Zika epidemic in the country (between 31/May/2015 and 01/October/2016).

SIVIGILA publicly provides surveillance data that come from each department’s Secretariat of Health. Probable and confirmed cases of Zika treated in public and private health facilities must be reported following the protocols of the National Institute of Health of Colombia ^16^. Complete case definitions for Zika are included in the Supporting Information Text 1.

### Environmental and sociodemographic data

The elevation (in meters) of each municipality (Figure S2A) was calculated for the centroid of the urban area using the packages sf ^37^ and elevatr ^38^ in R ^39^. The shapefile with the urban areas was obtained at the geoportal of the Colombian National Administrative Department of Statistics - *Departamento Administrativo Nacional de Estadística* (DANE) ^40^.

Other environmental data used in this work were previously organized and made publicly available by Siraj et al. ^41^. From the data by epidemiological week and municipality, we calculated the average Normalized Difference Vegetation Index (NDVI) for the entire study period for each municipality (Figure S2B). The NDVI quantifies vegetation greenness by using remote sensing technologies. In our study area, the NDVI ranged from 0.22 to 0.80. Areas with NDVI close to +1 have a higher possibility of being dense vegetation cover, while those with values close to 0 of being more urban with little vegetation cover. We obtained for each epidemiological week and municipality the maximum temperature in Celsius degrees (^°^C, Figures S2C and S3), the accumulated precipitation (in mm, Figures S2D and S4) and the relative humidity (in %, Figures S2E and S5).

The population estimates for 2015 and 2016 were obtained at the DANE website ^42^. We calculated the population density by dividing the population of each municipality by its area in km^2^ from a shapefile of the municipalities’ limits obtained at the DANE geoportal ^40^. Figure S6A shows the map with the mean population density (2015-2016) by km^2^ for each municipality.

Finally, we obtained the percentage of the population with unsatisfied basic needs (UBN) from the national census of 2018^43^ (Figure S6B). The UBN is an index provided by DANE that captures socioeconomic vulnerabilities such as inadequate housing conditions, overcrowded households, inadequate or no access to basic sanitation, children not attending school in the household, and household with elevated economic dependency ^44^.

### Statistical model

Let *y*_*it*_ denote the number of Zika cases reported in municipality *i* = 1, …, 1, 121 during week *t* = 1, …, 70. We assumed that Zika could be in one of three disease states within each municipality *i* during each week *t* denoted by the indicator *S*_*it*_,

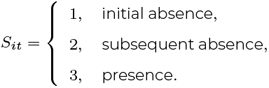

If at least one Zika case was reported, i.e. *y*_*it*_ *>* 0, we assumed the disease was present, i.e. *S*_*it*_ = 3. However, Zika may have been circulating undetected and as such, *S*_*it*_ was treated as an unknown parameter in the model and estimated whenever *y*_*it*_ = 0.

If Zika was present (*S*_*it*_ = 3), we assumed the reported cases were generated by a negative binomial distribution and if Zika was absent (*S*_*it*_ = 1 or 2), that no cases were reported,

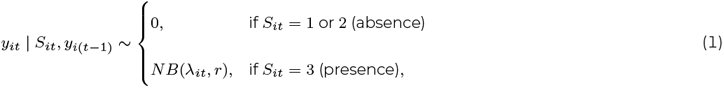

where *λ*_*it*_ is the expected number of reported cases given Zika was present and *r* is an overdispersion parameter so that the variance is given by 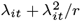.

To capture the transmission process of Zika when it was present, we assumed that *λ*_*it*_ follows an endemic/epidemic model ^45^,

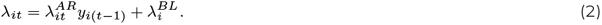

In (2), 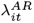 represents the transmission intensity rate of Zika when it was present which was assumed to depend on a vector of spatio-temporal covariates ***x***_*it*_ through a mixed log-linear regression,

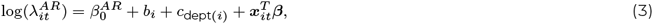

where *b*_*i*_ and *c*_dept(*i*)_ are zero mean municipality and department specific normal random intercepts. In (2), *λ*^*BL*^ is the baseline component representing the expected number of reported cases when no cases were reported in the previous week. We assumed *λ*^*BL*^ can vary between municipalities, so that, 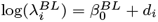, where *d*_*i*_ is a municipality specific random intercept.

As we would generally expect periods of Zika presence and absence to last several consecutive weeks, we used a Markov chain for modeling the switching between the states. Therefore, the current disease state in municipality *i S*_*it*_ only depends on the previous state *S*_*i*(*t−*1)_ and potentially on the covariates and random effects. The probabilities of transitioning from *S*_*i*(*t−*1)_ = *j* to *S*_*it*_ = *k* for *j, k* = 1, 2, 3 are given in the following within-area transition matrix,

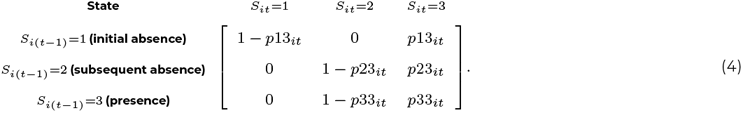

The probability of Zika emerging in municipality *i* during week *t, p*13_*it*_ in (4), was assumed to depend on a vector of space-time covariates ***z***_*it*_ through a mixed logistic regression,

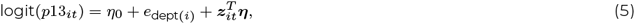

where *e*_dept(*i*)_ is a department (province) specific normal random intercept. As the disease states *S*_*it*_ and *S*_*i*(*t−*1)_ were not known when zero cases were reported, often the transition of (4) was not observable. Therefore, we did not include municipality specific random intercepts in the transition probabilities as it made model fitting unstable due to the amount of missing information. The probability of Zika re-emergence, *p*23_*it*_ in (4), was assumed to be equal to the probability of emergence plus a shift in intercept,

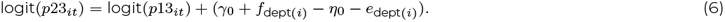

The shift in intercept was needed to account for the study start date. For example, if the study began two weeks earlier, the probability of emergence would reduce but the probability of re-emergence would not change. Also, as the disease established itself at some point in time, there may have been a shift in conditions not entirely accounted for by the covariates.

Finally, the probability of Zika persistence, *p*33_*it*_ in (4), was modeled similarly to emergence but the covariates, ***w***_*it*_, and their effects were allowed to differ,

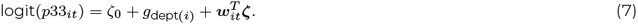

### Model specification

The relative humidity and the maximum temperature were included in the model lagged by one week to account for the time elapsed, on average, between an infected mosquito biting a person and the onset of symptoms ^46^. The weekly accumulated precipitation was lagged by four weeks to account for the additional time needed for a possible increase in the *Aedes* mosquitoes population after a rainy week.

All covariates were included in the emergence (5), re-emergence (6), and transmission intensity rate (3) equations. For the transmission intensity rate, we also considered the cumulative incidence of Zika up to four weeks prior and its square to account for the potentially non-linear effect of the depletion of the susceptible population on the transmission intensity rate. The four weeks lag was defined considering the average time elapsed between the person getting bitten by an infected mosquito and developing immunity against the Zika virus ^46–48^.

The largest factor affecting Zika persistence is likely the previous number of cases as the disease would continue to be present when there are many infectious individuals. Therefore, due to a high amount of multicollinearity, we only included the previous week’s cases and the population density as covariates potentially associated with Zika persistence.

The population density and the previous week cases were log transformed to reduce high skewness. All covariates were standardized to improve mixing of our Bayesian algorithm and to facilitate comparisons between covariate effect sizes.

### Model fitting

To fit the model [1]-(7), we took the Bayesian approach described in ^34^. When there were no cases reported by a municipality, the disease state of the model was not known as Zika could have been either absent, initially or subsequently, or present but undetected by the surveillance system. To help quantify uncertainty in the unknown disease states whenever zero cases were reported by a municipality, we calculated the posterior probability, i.e. the probability given all observed data, of Zika initial absence, presence, and subsequent absence ^34^.

An initial distribution for the disease state in week 1 was required to fit the model. If a municipality reported a positive number of cases in week 1, then we knew Zika was present, otherwise, we assumed there was a 5% chance Zika was present but undetected and a 95% chance Zika was initially absent.

We fitted the models using `NIMBLE` ^49–51^ in `R`^39^. Wide priors were assumed for all model parameters. Convergence was checked using the GelmanRubin statistic (all estimated parameters<1.02), the minimum effective sample size (>2000) and by visually examining the traceplots ^52^. For the maps and graphs, we used `ggplot2`^53^ and `colorspace`^54^ in `R`^39^. Codes are available in https://github.com/Dirk-Douwes-Schultz/Zika_Colombia_Markov_switching.

To check model fit, we assessed plots of fitted values compared to observed values. Broadly, the fitted values were constructed through simulation from the fitted model. See the Supporting Information Text 2 for more details about model fitting, estimation of the unknown disease states, and the fitted values.

### Ethical considerations

This study was approved by the Science and Health Research Ethics Committee (*Comité déthique de la recherche en sciences et en santé* - CERSES) of the University of Montreal, approval number CERSES-19-018-D.

## RESULTS

Between epidemiological weeks 22/2015 and 39/2016, there were 72,031 Zika cases reported to the Colombian national surveillance system. The first cases were residents of three municipalities located in the department of Norte de Santander (San José de Cúcuta, El Zulia and Puerto Santander), at EW 26/2015. Valle del Cauca, Norte de Santander, and Santander were the departments with the highest number of reported cases (20,965, 8,666 and 8,659, respectively) (Figure S7). During the study period, 348 (31%) municipalities did not reported Zika cases (Table S1). The departments with higher percentage of municipalities not reporting Zika cases were Guainía, Vaupés, Nariño, Chocó, Amazonas and Boyacá. Higher cumulative incidences per 10,000 inhabitants were observed in municipalities south of the Colombian Andes and in the departments of Norte de Santander, Valle del Cauca and Archipelago of San Andrés, Providencia, and Santa Catalina (Figure S8).

### Zika association with covariates

Figure 4A shows the posterior distribution of the transmission intensity rate ratio (TRR) and odds ratio (OR) associated with the covariates in standardized form.

**Figure 4:**
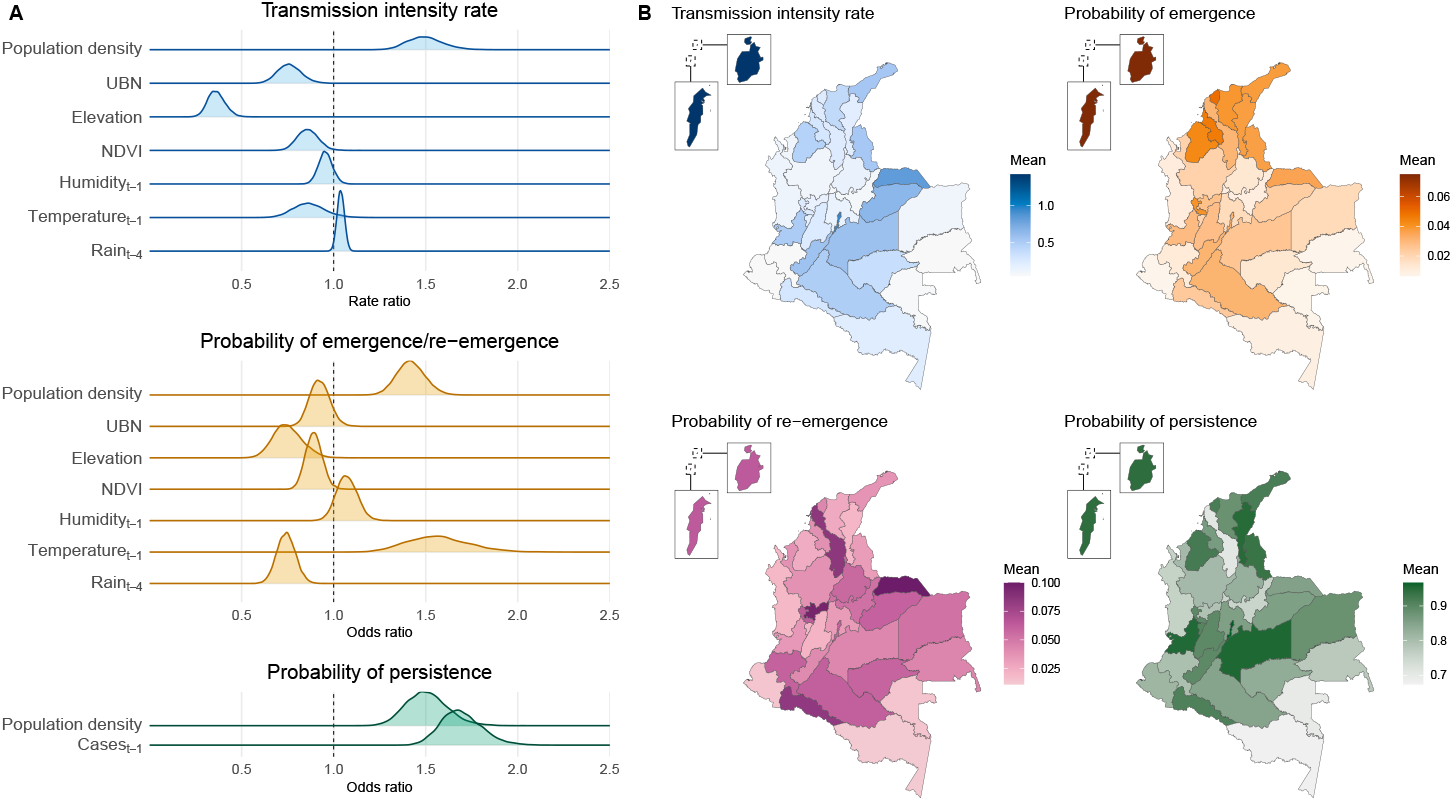
A) Posterior distribution of the rate ratio/odds ratio associated with standardized covariates* and the transmission intensity rate, probabilities of emergence and re-emergence, and probability of persistence of Zika, and B) posterior mean of the transmission intensity rate, probability of emergence, re-emergence, and persistence of Zika by department after adjusting for the department-specific random effect and the average values of the covariates in the department, epidemiological weeks 22/2015 to 39/2016, Colombia. *Population density and cases*t−*1 are log transformed. UBN = percentage of the population with unsatisfied basic needs. NDVI = Normalized Difference Vegetation Index.

Assuming a 95% credible interval (CI), when Zika was present, transmission was more intense in areas with higher population density (TRR mean 1.49, 95%CI 1.33;1.70), lower percentage of the population with unsatisfied basic needs (TRR 0.76, 95%CI 0.64;0.89), lower altitude (TRR 0.36, 95%CI 0.28;0.46), and/or less vegetation cover (TRR 0.86, 95%CI 0.75;0.98). It is worth mentioning that the transmission intensity rate showed a borderline direct association with the weekly accumulated rain (TRR 1.04, 95%CI 1.00;1.08, lagged by four weeks).

Figure S9 shows the association between cumulative incidence (lagged by four weeks) and the Zika transmission intensity rate by department after adjusting for the department-specific random effect and the average values of the covariates in the department. A rapid decrease in the transmission intensity rate was estimated with the increase of the cumulative incidence up to 5 cases per 10,000 inhabitants, more apparent for the department of Archipelago of San Andrés, Providencia and Santa Catalina. Above 5 cases per 10,000 inhabitants, the transmission intensity rate slightly decreases with the increase of the cumulative incidence.

Zika emerged and re-emerged sooner in municipalities with lower altitude (OR mean 0.75, 95%CI 0.61;0.91), less vegetation cover (OR 0.89, 95%CI 0.81;0.98) and/or less weekly accumulated rain (OR 0.74, 95%CI 0.64;0.84, lagged by four weeks). Zika also emerged and re-emerged sooner in more densely populated municipalities (OR 1.42, 95%CI 1.28;1.58) and/or with higher weekly maximum temperatures (OR 1.56, 95%CI 1.27;1.92, lagged by one week).

The disease persisted for longer in more densely populated municipalities (OR mean 1.50, 95%CI 1.30;1.74). Controlled by the population density, Zika was more likely to persist in a municipality when and where a higher number of cases was reported in the previous week (OR 1.69, 95%CI 1.50;1.94).

### Department-specific transmission intensity rate and probabilities of emergence, re-emergence, and persistence

After adjusting for the department-specific random effects and average values of the covariates in each department, higher transmission intensity rates were estimated in the Archipelago of San Andrés, Providencia and Santa Catalina, followed by departments located south of the Colombian Andes, Valle del Cauca, Norte de Santander, and in the north of the country (Figures 4B and S10). A similar spatial pattern was observed for disease emergence, but departments in the north of the country showed, on average, a higher probability of emergence compared to departments south of the Andes. However, it is worth mentioning that the differences in the probability of emergence across the departments were very small (Figure S11). The same is true for the probability of re-emergence. Zika was more likely to persist in the departments of Archipelago of San Andrés, Providencia and Santa Catalina, Meta, Bogotá D.C, Valle del Cauca, Córdoba, Cesar, Norte de Santander, La Guajira, and Putumayo (Figures 4B and S11).

### Zika transmission intensity rate

Maps showing the evolution of the transmission intensity rate over the entire study period are displayed in Movie S1. The transmission intensity rate generally declines over the study period as the susceptible population becomes depleted. The effects of the covariates can be clearly seen in the spatial variation of the maps as municipalities located in the mountains typically experience much lower transmission of Zika. Additionally, heterogeneity introduced by the random effects is also clearly visible. For example, there are certain municipalities on the Pacific coast (in the departments of Chocó, Cauca, and Nariño) with very little transmission rates of Zika despite being at a low elevation. At the beginning of the study period, the highest Zika transmission rates were observed in the municipalities of San Andrés (Archipelago of San Andrés, Providencia and Santa Catalina department), Guadalajara de Buga (Valle del Cauca), Tauramena (Casanare), Bucaramanga (Santander), and Saravena (Arauca).

Figure S12 depicts the posterior mean of the baseline expected number of reported cases by municipality, representing the expected number of reported cases in a given week if no cases were reported in the previous week. It ranged from 0.13 in González (Cesar department) to 2.86 in Chaguaní (Cundinamarca department). Higher baselines were mainly observed in some municipalities from the departments of Cundinamarca, Magdalena, Norte de Santander, Valle del Cauca, Córdoba, Tolima, Antioquia, Casanare, and Santander.

### Fitted values and posterior probability of Zika presence

For Figure 5, we selected the first three municipalities from Figure 1 as examples to compare the observed number to the fitted number of Zika cases (first row) and the estimated posterior probabilities of each disease state (second row).

**Figure 5:**
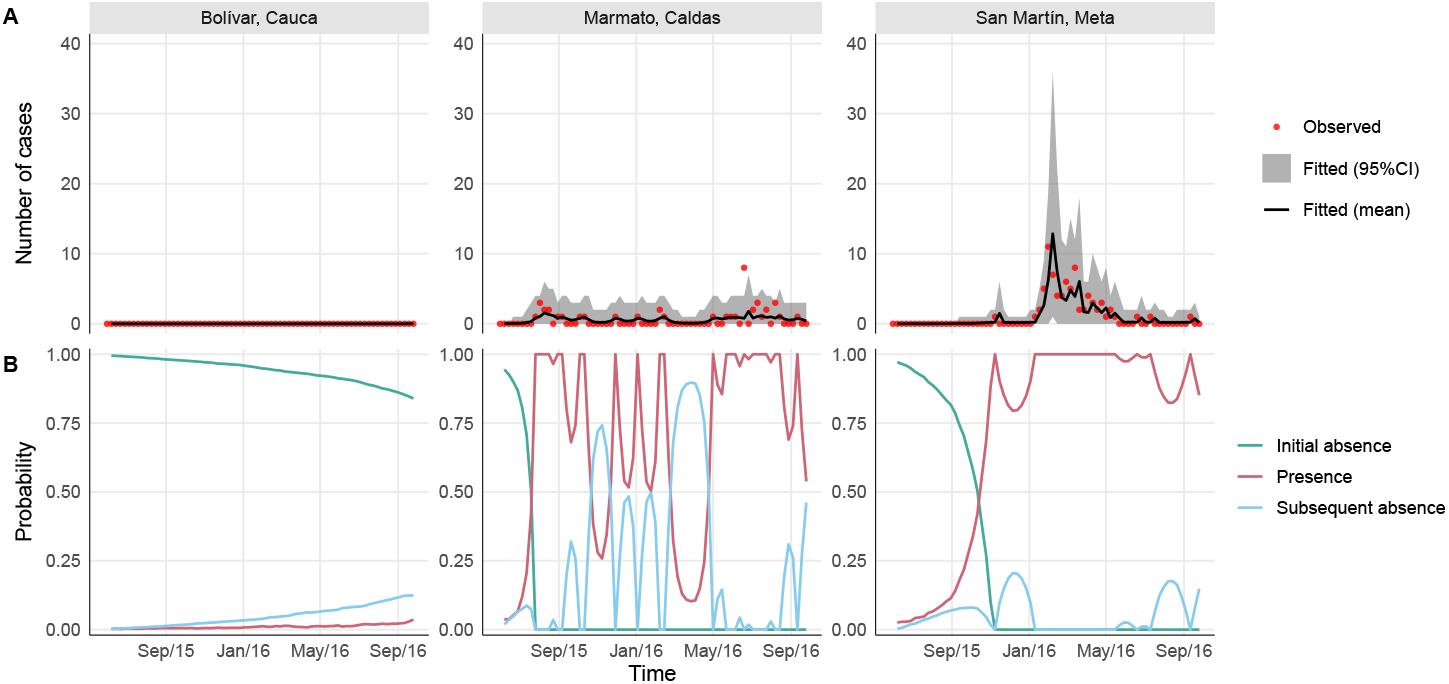
A) Observed versus fitted number of Zika cases (mean and 95% Credible Interval - CI) and B) posterior probability of initial absence, presence, and subsequent absence of Zika by epidemiological week (EW) in three selected municipalities, EWs 22/2015 to 39/2016, Colombia.

As can be seen in the first row of Figure 5, the fitted model was able to reproduce the reported number of cases well. The fitted mean follows the observed trend and the 95% credible intervals capture the majority of the observations. Note for the second row of Figure 5, that when cases were reported by a municipality, the posterior probability of Zika being present was always 1. Otherwise, if zero cases were reported, the disease could have either been absent, initially (probability given by green line) or subsequently (blue line), or present but undetected (red line). In Marmato and San Martín, the posterior probability of Zika presence began to increase noticeably several weeks before the first reported cases and the posterior probability remained high when no cases were reported afterwards. In general, as the number of consecutive weeks with no cases being reported increased, the model becomes more certain that Zika is truly absent and not undetected. In Bolívar, which never reported any cases, the model estimated a 15% chance that Zika was no longer in the initial absence state in the last week, implying it may have circulated undetected at some earlier point in time. These patterns in the posterior probabilities were typical across the other municipalities.

In Figure 6, we compared the maps with the posterior probability of Zika presence (first row) with the maps with the observed number of reported cases (second row) at four different moments during the study period. The maps for the entire study period are displayed in Movie S2. When there are reported cases, the probability of Zika presence was always 1. However, we observed that the estimated probability of Zika presence increased before cases began to be reported, and remained high in certain municipalities, some of which had no reported cases of Zika. This was partly due to the department specific random intercepts in the model, as municipalities with no cases would have an increased probability of Zika presence if other municipalities in the same department reported cases.

**Figure 6:**
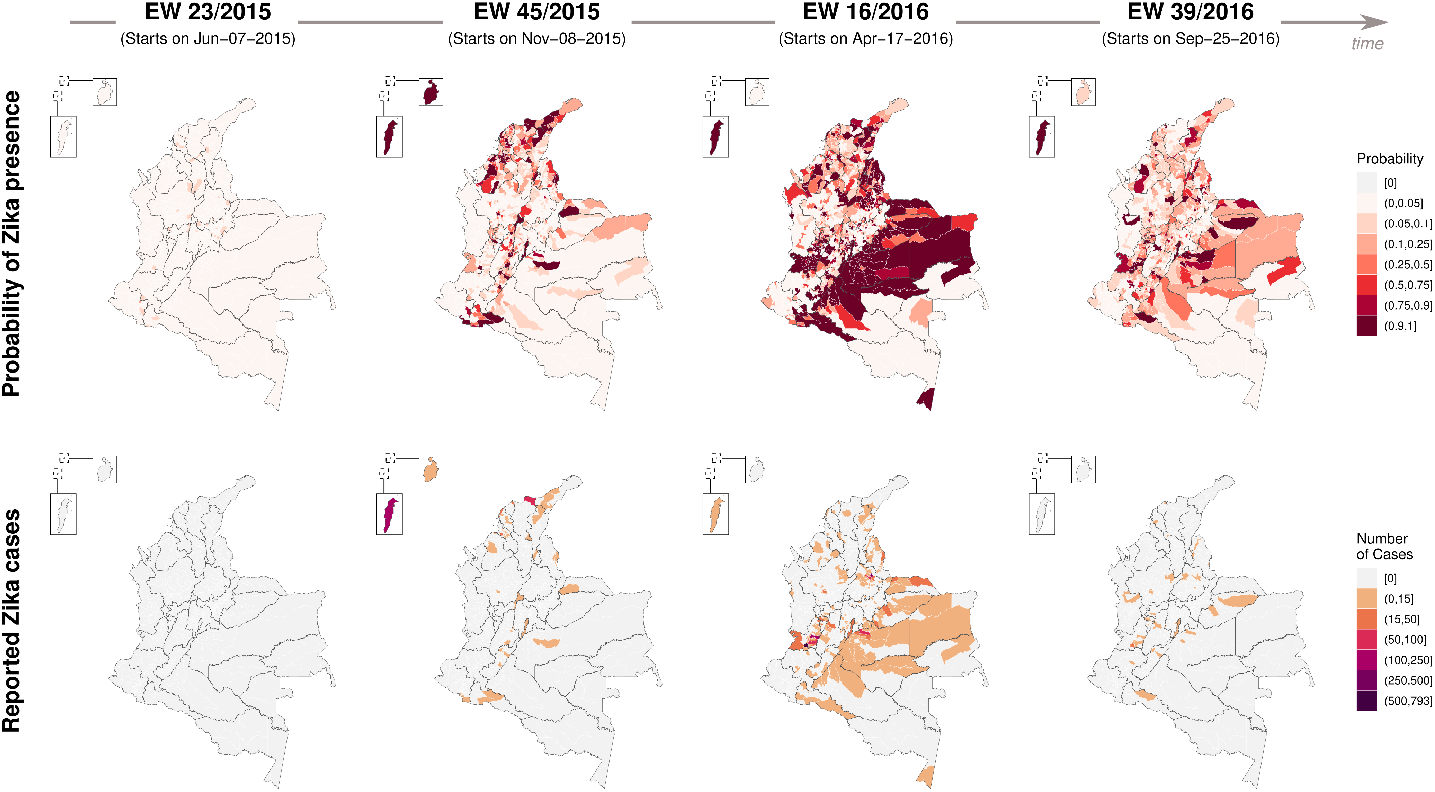
Posterior probability of Zika presence (first row) and observed number of reported Zika cases (second row) by municipality in four moments of time between epidemiological weeks (EWs) 22/2015 and 39/2016, Colombia.

## DISCUSSION

In this work, we studied the first Zika epidemic in Colombia using a novel modeling approach, which could manage the excessive amount of zeros in the data as well as the different temporal patterns of reported cases between the municipalities. The proposed model estimated the association of environmental and sociodemographic covariates with the probabilities of emergence, re-emergence, and persistence, and with the transmission intensity rate of Zika. Our results suggest that the population density, elevation, and maximum temperature were the main contributors of the first Zika epidemic in Colombia. We were also able to estimate the probability of Zika presence by week and municipality, which often increased before the first official report of Zika.

Population density had a strong positive association with the transmission intensity rate, the probabilities of emergence, re-emergence, and persistence of Zika (Figure 4A). Although an emerging virus may enter a territory through a smaller city, it will likely establish itself more readily in larger cities with higher population densities, from where it may spread to the rest of the territory ^15^. Densely populated areas provide more opportunities for the virus to infect vectors and humans.

We found an inverse association between the percentage of the population with unsatisfied basic needs and the Zika transmission intensity rate, and the direction of this association needs to be interpreted with caution. The majority of municipalities with higher percentages of their populations with unsatisfied basic needs belong to the following departments: Chocó, Nariño, Vichada, Guainía, Vaupés, Amazonas, and La Guajira (Figure S6B). Among the 138 municipalities of these departments, 95 (or 68.8%) did not report any cases of Zika during the study period (Figure S8). These particular locations are known to have problems of underreporting and the local populations face important barriers to access the health system. These can be explained by the presence of extensive jungle-covered areas, an ongoing violent conflict, a limited number of healthcare facilities, and potentially weaker surveillance systems compared to other areas. Nonetheless, the inverse association found could be a consequence of the spatial unit size used in the analysis. In Colombia, there are important social inequalities that cannot be fully captured by a single socioeconomic index value for an entire municipality. To better understand the role of socioeconomic conditions in the Zika epidemic, it may be necessary to work with smaller spatial units in order to have sufficient resolution to capture these inequalities.

Zika’s transmission intensity rate, probability of emergence and probability of re-emergence were higher in more urban municipalities, i.e., with less vegetation cover (represented by the NDVI), and/or with lower altitude (Figure 4A). The inverse association with NDVI may be explained by the preference of *Aedes aegypti* for urban settings ^55^, and also by the presence of heat islands in urban settings affecting the microclimate. The estimated effect of elevation and temperature on the transmission intensity rate needs to be interpreted with caution. It should not be concluded that there is no association between temperature and transmission intensity, only that, after accounting for elevation, the temperature effect was very small towards null. As can be seen in Figure S2 panels A and C, temperature and elevation had a strong inverse correlation, with areas with lower temperatures mostly found at higher elevations. Therefore, areas with low temperatures did experience lower transmission of Zika on average. Also, the temperature in Colombia remained relatively constant over time (Figure S3), mainly varying spatially. Therefore, it is possible that elevation was capturing part of the association between the Zika transmission intensity rate and temperature.

Climate factors (temperature and rain) were strongly associated with the emergence and re-emergence of Zika but less so with its transmission intensity rate (Figure 4A). In addition to the possible explanations discussed above, it is possible that there were interactions between different climate factors (including but not limited to temperature, humidity and rain) that would require more complex modeling structures to be captured. We hypothesize that climate factors, along with elevation and NDVI, captured the presence of the *Aedes aegypti* mosquitoes, which is necessary for local transmission to occur. Using climate data as proxies of the mosquitoes’ presence is advantageous, as such data tend to be more readily available than mosquito data.

An inverse association between rain and the probabilities of Zika emergence and re-emergence was found. During the study period (2015-2026), the El Niño event was affecting South America, causing warmer temperatures and droughts in Colombia and boosting the transmission of *Aedes*-borne diseases ^56,57^. A negative association between precipitation and the number of dengue cases was found in a previous study evaluating the effects of local climate and El Niño in Colombia ^57^. Analyzing data by department, Chien et al. ^26^ found both positive and negative associations between rain and the risk of Zika in Colombia. Although rain may increase mosquito density by creating potential breeding sites, heavy rain can wash away the eggs and larvae. Also, droughts may result in people storing water in improvised containers inside their households as a response to water supply interruptions. These improvised water-filled containers are well-suited for *Aedes aegypti* breeding and may favor an increase in mosquito density. An association between periods of drought and increased risk of *Aedes*-borne diseases has been previously described ^5,58^.

The increase in the probability of disease presence weeks before the first reported cases, indicates that Zika circulated undetected in the early phases of the epidemic. This is expected, considering that Zika was a new emerging disease and there was no active surveillance implemented to detect the entry of the virus into the country. The model presented in this study has the potential to be adapted for similar scenarios and provide insight to inform our understanding of emerging and re-emerging diseases’ spatio-temporal distributions and, ultimately, help guide control and prevention strategies. The results may also indicate locations with notifiable disease reporting issues when a high probability of disease presence is estimated but when no cases are reported.

We recommend the implementation of effective measures, including enhanced surveillance and vector-control strategies, in Colombian urban centers, particularly those with high population density. These measures are essential for mitigating the introduction of other emerging viruses transmitted by *Aedes*, as well as reducing the burden of the *Aedes*-borne diseases that are already endemic in the country. On the other hand, the processes of urbanization, resulting in increased population density and loss of vegetation cover, increase the risk of *Aedes*-transmitted diseases. Therefore, strategies for sustainable human development that prioritize the preservation and augmentation of vegetation cover should be considered in order to effectively combat these diseases. Additionally, ensuring access to piped water without supply interruptions is crucial. When that is not feasible, the population should also be educated and provided with means to correctly store water to prevent the creating of breeding sites for the mosquitoes.

This study has limitations. Despite the SIVIGILA protocol foreseeing active surveillance in certain situations, most Zika reported cases come from passive surveillance ^16^. Hence, our study population included mostly patients who developed symptoms and sought health care. Also, the observed counts may be underestimated. Underreporting is a common limitation when working with surveillance data and has been detected in Colombia’s surveillance system ^59^. The majority of cases were confirmed by clinical and epidemiological criteria, with only a small proportion being laboratory-confirmed. We aggregated the data based on municipality of residence, although cases may have been infected elsewhere. The climate factors were included in the model with predefined lags, although we acknowledge that more complex structures considering different lags, interactions, and possibly nonlinear effects may be necessary to better capture the association between climate and *Aedes*-borne diseases ^60^. Including such structures in the scope of the present work is challenging, considering the large number of model parameters, areas, and time points.

The range of the *Aedes aegypti* mosquito has been expanding as a consequence of global warming ^61^. This is resulting in increased occurrence of *Aedes*-transmitted diseases in endemic areas but also the emergence in previously unaffected areas ^62^. Large epidemics of Zika can occur in areas where most of the population was never exposed to the virus. Although Zika is usually mild, infection during pregnancy can cause congenital malformation in the fetus and its risk should not be neglected ^63^. Our model can be used to better understand the factors contributing to disease emergence, re-emergence, persistence, and transmission intensity at high spatial and temporal resolution. This can provide valuable insights into the characteristics of areas that should be prioritized for interventions such as vector-control measures, enhanced surveillance, and public health campaigns.

## Data Availability

The data used in this study are secondary data and are publicly available. The data on Zika cases can be downloaded at the SIVIGILA website (http://portalsivigila.ins.gov.co/). Population data and the percentage of people with unsatisfied basic needs can be found at the DANE website (https://www.dane.gov.co/). Environmental data was organized and made available by Siraj et al. (https://doi.org/10.5061/dryad.83nj1). Processed data and codes are available in https://github.com/Dirk-Douwes-schultz/Zika_Colombia_Markov_switching.

## DATA AVAILABILITY

The data used in this study are secondary data and are publicly available. The data on Zika cases can be downloaded at the SIVIGILA website (http://portalsivigila.ins.gov.co/). Population data and the percentage of people with unsatisfied basic needs can be found at the DANE website (https://www.dane.gov.co/). Environmental data was organized and made available by Siraj et al. ^64^ (https://doi.org/10.5061/dryad.83nj1). Processed data and codes are available in https://github.com/Dirk-Douwes-Schultz/Zika_Colombia_Markov_switching.

## ACKNOWLEDGMENTS

The authors would like to thank the healthcare workers who collect the surveillance data from the patients and report the cases, and the National Institute of Health of Colombia for making the diseases’ surveillance data publicly available. We also would like to thank Dr. Siraj and collaborators for processing and making the environmental data available. This work was supported by the Canadian Institutes of Health Research (CIHR), grant number 428107. Douwes-Schultz is grateful for financial support from IVADO and the Canada First Research Excellence Fund/Apogée (PhD Excellence Scholarship 2021-9070375349). Schmidt acknowledges nancial support from the Natural Sciences and Engineering Research Council (NSERC) of Canada (Discovery Grants RGPIN-2017-04999). The funders had no role in study design, data collection and analysis, decision to publish, or preparation of the manuscript.

## AUTHOR CONTRIBUTIONS

L.P.F., D.D.S, K.Z., A.M.S., M.C., G.I.J.R., C.G.B., B.N.R. conceptualized research. L.P.F. and D.D.S. designed research, performed research, analyzed data and wrote the paper. A.M.S., B.A.M., J.E.S.F., C.G.B., B.N.R., G.I.J.R., M.C. and K.Z. reviewed and approved the paper.

