## Supporting information for "Zika epidemic in Colombia and environmental and sociodemographic contributors: an application of a space-time Markov switching model"

**Laís Picinini Freitas [a,b,1*] & Dirk Douwes-Schultz [c,1*], Alexandra M. Schmidt [c], Brayan Ávila Monsalve [d], Jorge Emilio Salazar Flórez [e], César García-Balaguera [d], Berta N. Restrepo [e], Gloria I. Jaramillo-Ramirez [d], Mabel Carabali [c], Kate Zinszer [a,b]**
[a] École de Santé Publique, Université de Montréal, Montreal, Canada
[b] Centre de Recherche en Santé Publique, Montreal, Canada
[c] Department of Epidemiology, Biostatistics and Occupational Health, McGill University, Montreal, Canada
[d] Faculty of Medicine, Universidad Cooperativa de Colombia, Villavicencio, Colombia
[e] Instituto Colombiano de Medicina Tropical, Universidad CES, Medellín, Colombia
[1] L.P.F. and D.D.S. contributed equally to this work.

### 1. ZIKA CASE DEFINITION

The following criteria describe the case definition for Zika cases adopted by the Colombian National Institute of Health [1].

#### Probable case

Patient with a rash and one or more of the following symptoms not explained by other medical conditions: fever not greater than 38.5°C, nonpurulent conjunctivitis or conjunctival hyperemia, arthralgia, myalgia, headache, or malaise. Additionally, one of the following conditions: i) Person who visited, two weeks before the onset of symptoms, countries or municipalities located between 0 and 2,200 m above sea level, with or without confirmed indigenous circulation of the Zika virus; ii) Person who had sexual contact without barrier protection two weeks before the onset of symptoms with a person who in the eight weeks prior to sexual contact visited areas with confirmed Zika transmission and/or areas with the presence of *Aedes* mosquitoes.

#### Confirmed case by clinical epidemiological criteria

Patient who had been in countries or municipalities located between 0 and 2,200 meters above sea level with confirmed autochthonous circulation of the Zika virus two weeks before the onset of symptoms and who presented a rash and one or more of the following symptoms that were not explained by other medical conditions: fever not higher than 38.5 °C, non-purulent conjunctivitis or conjunctival hyperemia, arthralgia, myalgia, headache or general malaise.

#### Confirmed case by laboratory

Case that met the definition for probable case and that presented a positive result for Zika virus by RT-PCR (or immunohistochemistry in histopathological analysis) performed at the National Reference Laboratory of the National Institute of Health (INS) or collaborating centers designated by the INS.

### 2. ADDITIONAL INFORMATION REGARDING MODEL FITTING, ESTIMATION OF THE UNKNOWN DISEASE STATES AND THE FITTED VALUES

We fitted the model using a hybrid Gibbs sampler, described in more detail in [2], which we implemented in the R package `NIMBLE` [3]. Gibbs sampling begins by partitioning the unknown parameter vector into blocks and setting suitable random initial values for each block. Then the algorithm alternates between sampling each block of unknown parameters conditional on all other blocks and the observed data [4]. After an initial burn-in period the Gibbs sampler draws correlated samples from the joint posterior distribution, i.e., distribution given the observed data, of all unknown parameters. Although, it is possible for the samples to be too correlated to draw reasonable inference from and the Gibbs sampler can get stuck in local modes, therefore, it is important to assess convergence of the algorithm [5]. Note that the unknown state indicators, that is $S_{it}$ for all $i$ and $t$ such that $y_{it} = 0$, are a part of the unknown parameter vector in our application as the disease state is not known when no cases are reported. We sampled each unknown parameter, which was not an unknown state indicator and did not have a conjugate prior, individually, using an adaptive random walk Metropolis step [6]. We sampled all the unknown state indicators in a municipality jointly using a forward filtering backwards sampling (FFBS) algorithm [7]. Sampling the unknown state indicators of a Markov switch model jointly is typically much more efficient than sampling them individually [8].

Once a sample from the joint posterior has been obtained, the posterior probability that the disease was in disease state $s$, for $s = 1$ (initial absence), $s = 2$ (subsequent absence) and $s = 3$ (presence), at time $t$ in area $i$, assuming $y_{it} = 0$ so no cases were reported, is given by,

$$P(S_{it} = s|\boldsymbol{y}) \approx \frac{1}{Q-M} \sum_{m=M+1}^{Q} I[S_{it}^{[m]} = s],$$

where the superscript $[m]$ denotes a draw from the posterior distribution of the variable, $M$ is the size of the burn-in sample, $Q$ is the total number of iterations of the Gibbs sampler, $I[\bullet]$ is an indicator function and $\boldsymbol{y}$ is the vector of all observed data. In Figure 5B we plot $P(S_{it} = s|\boldsymbol{y})$ versus $t$ for each $s = 1, 2, 3$, and note $P(S_{it} = 3|\boldsymbol{y}) = 1$ if $y_{it} > 0$ as we know the disease is present. In Figure 6 (top) the maps show $P(S_{it} = 3|\boldsymbol{y})$ across $i$ for various times, note $P(S_{it} = 3|\boldsymbol{y})$ when $y_{it} = 0$ gives the probability, given all observed data, that the disease is present and thus circulating undetected in area $i$ at time $t$.

Finally, for the fitted values, we assumed a hypothetical new count $y^*_{it}$ that is generated by the model assuming the same disease state, past counts and parameters that generated $y_{it}$. We can then draw from the posterior of the fitted value $y^{*[m]}_{it} \sim p(y^*_{it}|\boldsymbol{y})$ by drawing $y^{*[m]}_{it}$ from a $NB(\lambda^{[m]}_{it}, r^{[m]})$ if $S^{[m]}_{it} = 3$ and setting $y^{*[m]}_{it} = 0$ otherwise, for $m = M+1, \ldots, Q$. Figure 5A shows the posterior mean and 95% credible interval of $y^*_{it}$, approximated by such draws, against the observed value $y_{it}$, versus $t$. We compared the simulated values from the model with the observed values to assess the fit of the model, which is common in Bayesian statistics and is often referred to as a "posterior predictive check" (see [4] Section 5.6 for instance).

**Movie S1.** Space-time distribution of the estimated Zika transmission intensity rate by municipality and epidemiological week (EW), EWs 23/2015 to 39/2016, Colombia.

**Movie S2.** Space-time distribution of the estimated probability of Zika presence and of reported Zika cases by municipality and epidemiological week (EW), EWs 23/2015 to 39/2016, Colombia.

**Table S1:** Number and percentage of municipalities by percentage of weeks with no reported Zika cases, EWs 22/2015 to 39/2016, Colombia.

| Percentage of weeks with no reported cases | Municipalities | |
|---|---|---|
| | N | % |
| [0] | 16 | 1.43 |
| (0-25) | 2 | 0.18 |
| [25-50) | 29 | 2.59 |
| [50-75) | 107 | 9.54 |
| [75-100) | 619 | 55.22 |
| [100] | 348 | 31.04 |
| | 1121 | 100.00 |

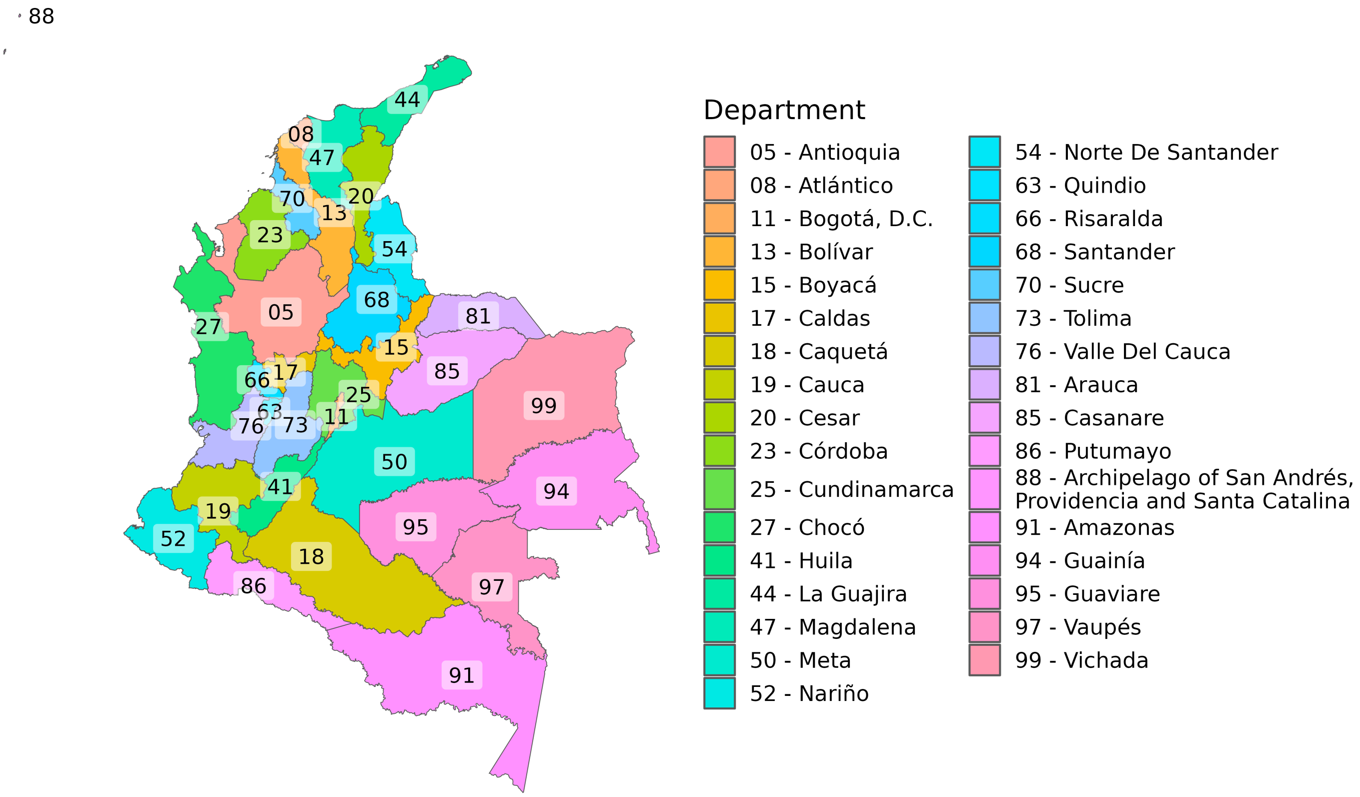


**Figure S1:** Departments of Colombia. Data source: Colombian National Administrative Department of Statistics - *Departamento Administrativo Nacional de Estadística* (DANE).

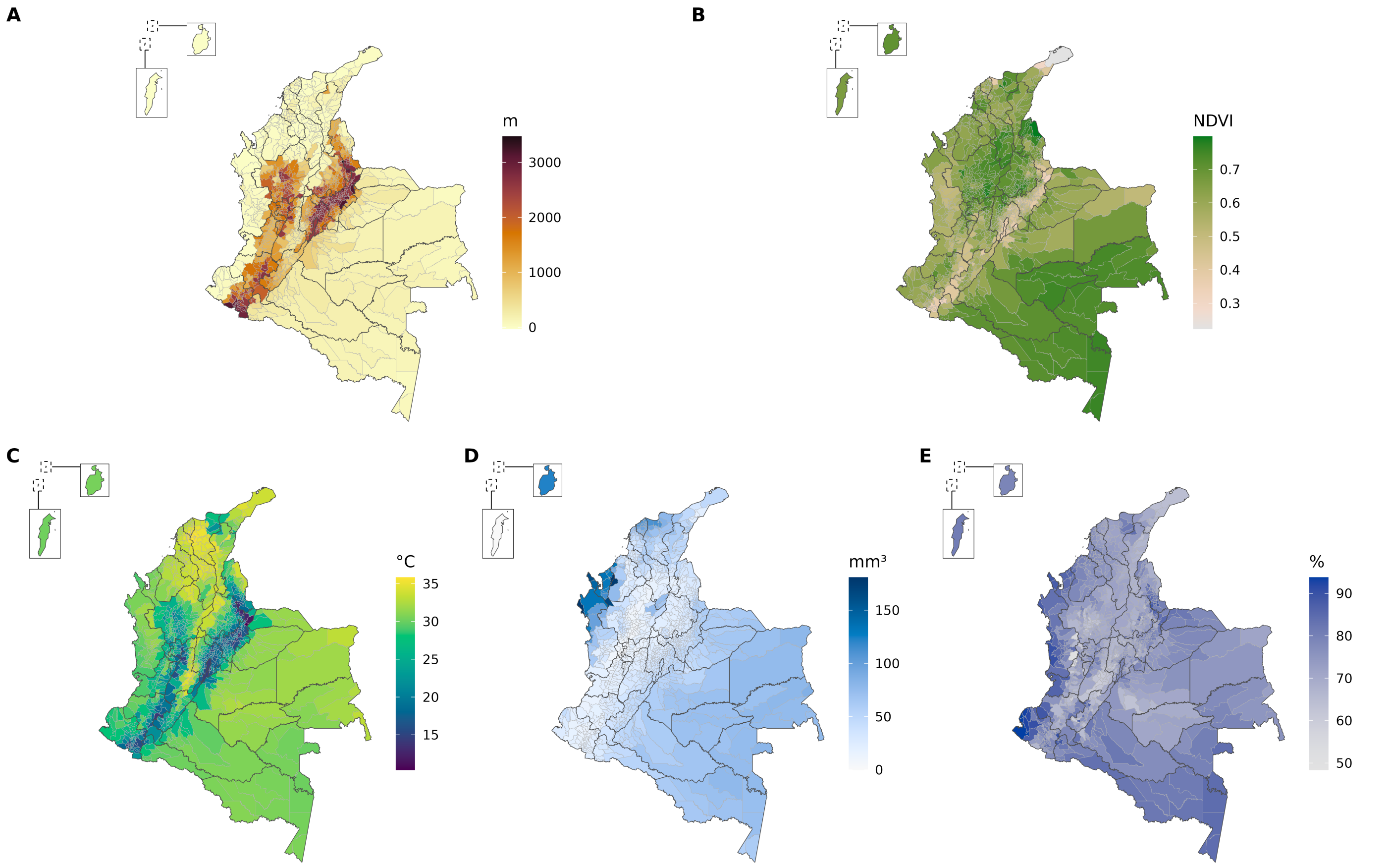


**Figure S2:** (A) Elevation, (B) mean Normalized Difference Vegetation Index (NDVI), (C) mean maximum temperature, (D) total accumulated precipitation, and (E) mean relative humidity by municipality, epidemiological weeks 22/2015 to 39/2016, Colombia. Data sources: Colombian National Administrative Department of Statistics - *Departamento Administrativo Nacional de Estadística* (DANE); Siraj et al. (2019) [9].

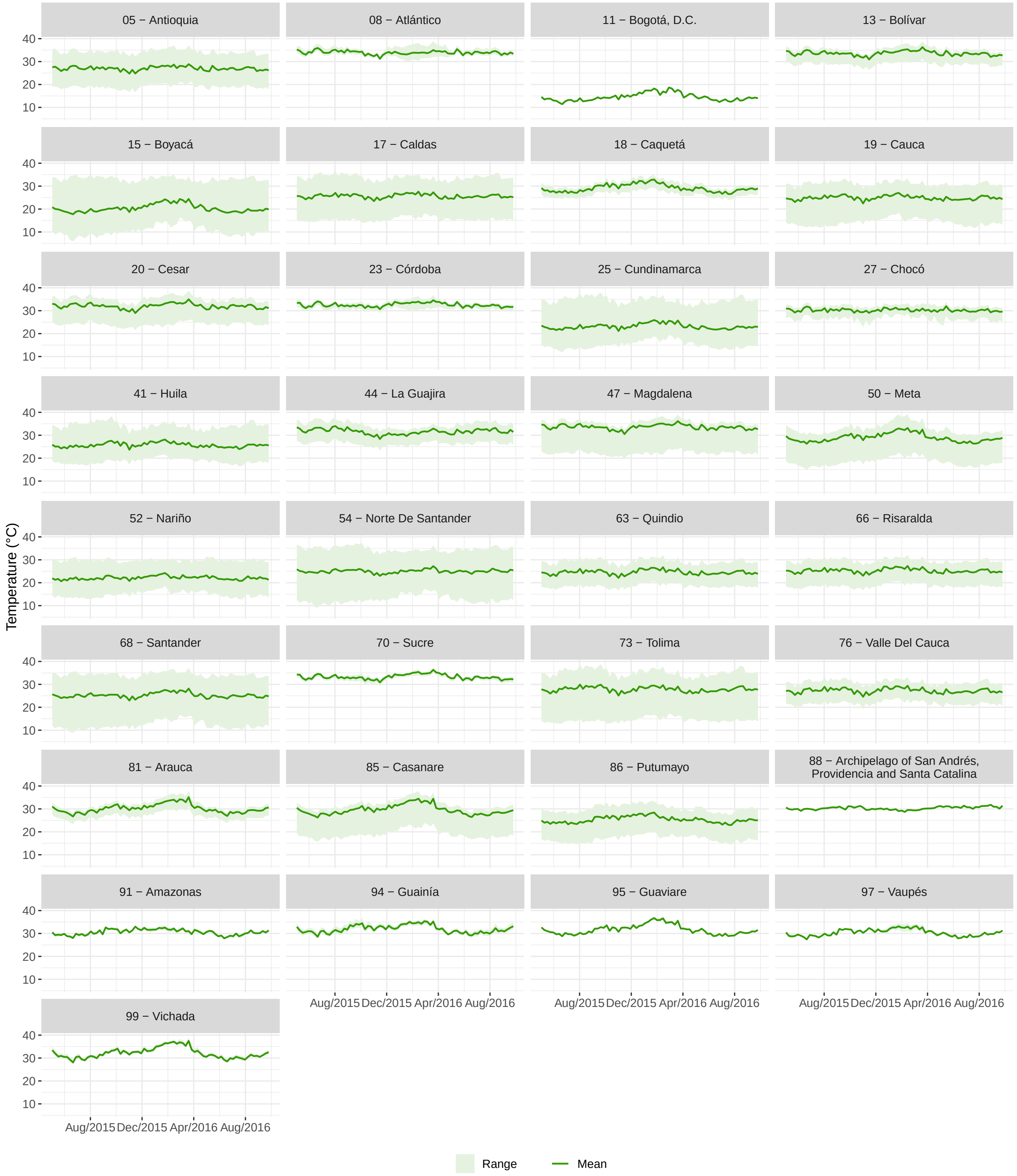


**Figure S3:** Maximum temperature (in °C) by department and epidemiological week (EW), EWs 18/2015 to 39/2016, Colombia. From the data by municipality, we calculated the mean and the range (minimum and maximum values) for each department. Data source: Siraj et al. (2019) [9].

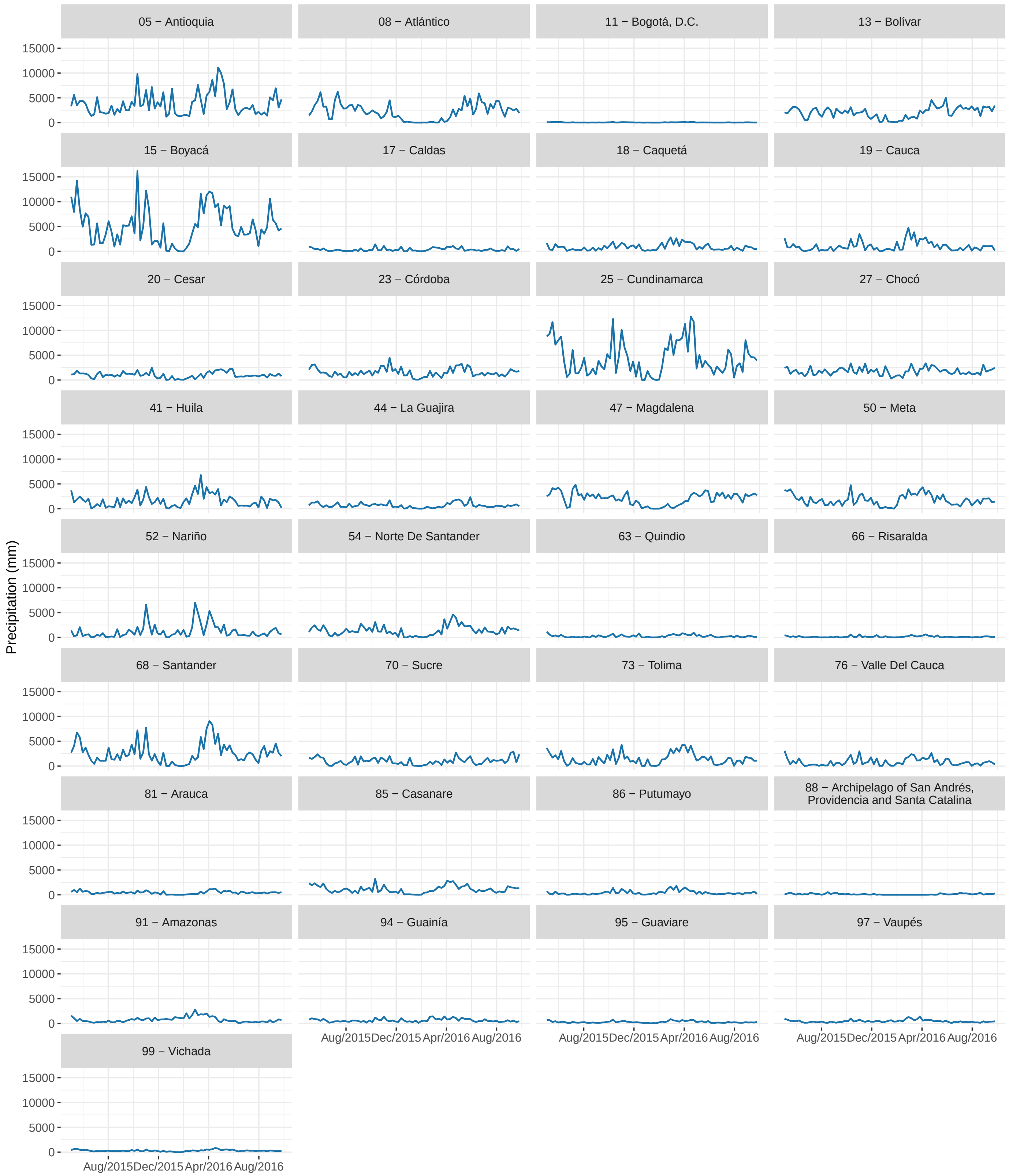


**Figure S4:** Accumulated weekly precipitation (in mm) by department and epidemiological week (EW), EWs 18/2015 to 39/2016, Colombia. The precipitation for each department was calculated by summing the data by municipality. Data source: Siraj et al. (2019) [9].

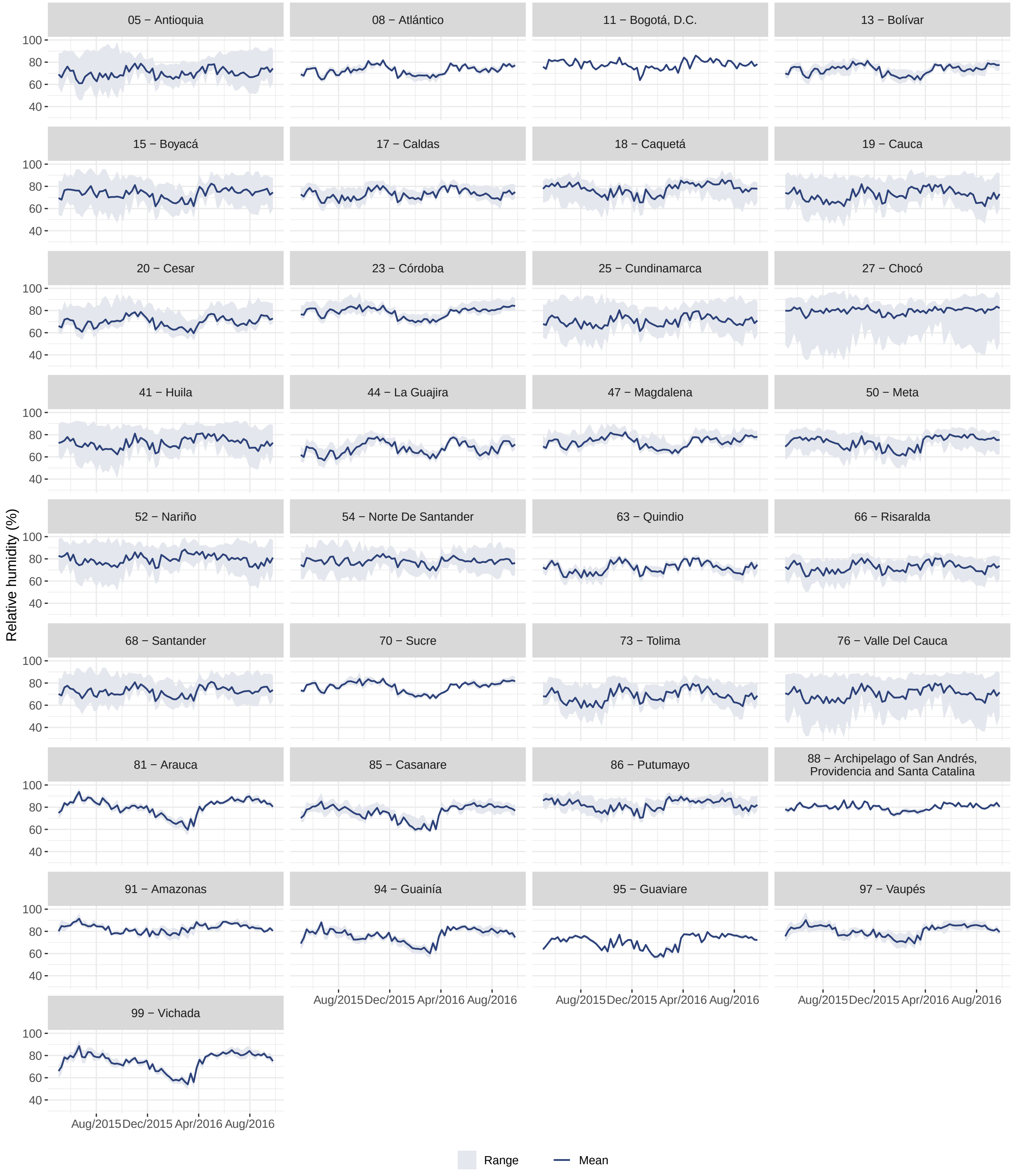


**Figure S5:** Relative humidity (%) by department and epidemiological week (EW), EWs 18/2015 to 39/2016, Colombia. From the data by municipality, we calculated the mean and the range (minimum and maximum values) for each department. Data source: Siraj et al. (2019) [9].

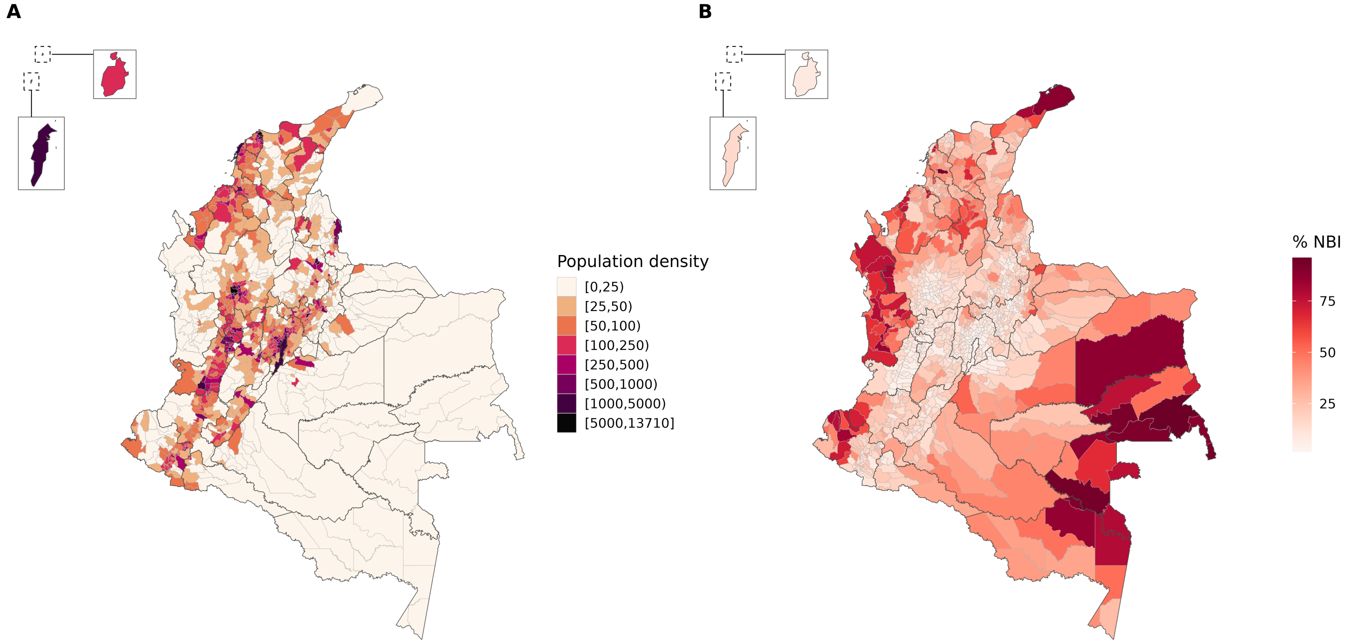


**Figure S6:** (A) Mean population density (2015-2016) by km² and (B) the percentage of population with unsatisfied basic needs (*Necesidades Básicas Insatisfechas* - NBI) (2018) by municipality, Colombia. Data source: Colombian National Administrative Department of Statistics - *Departamento Administrativo Nacional de Estadística* (DANE).

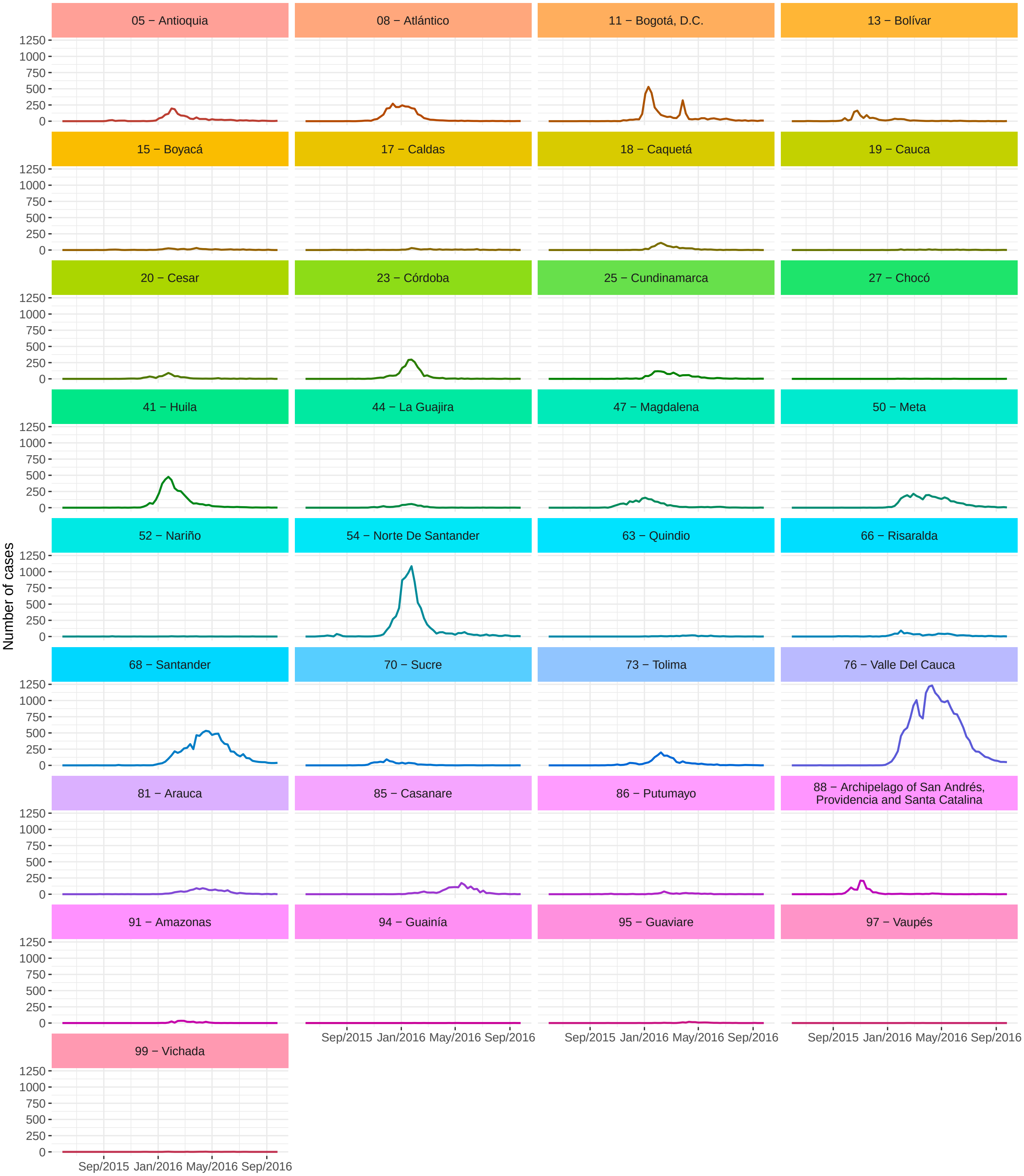


**Figure S7:** Number of reported Zika cases by department of residence and epidemiological week (EW) of first symptoms, EWs 22/2015 to 39/2016, Colombia. Data source: Colombian National Public Health Surveillance System - *Sistema Nacional de Vigilancia en Salud Pública* (SIVIGILA).

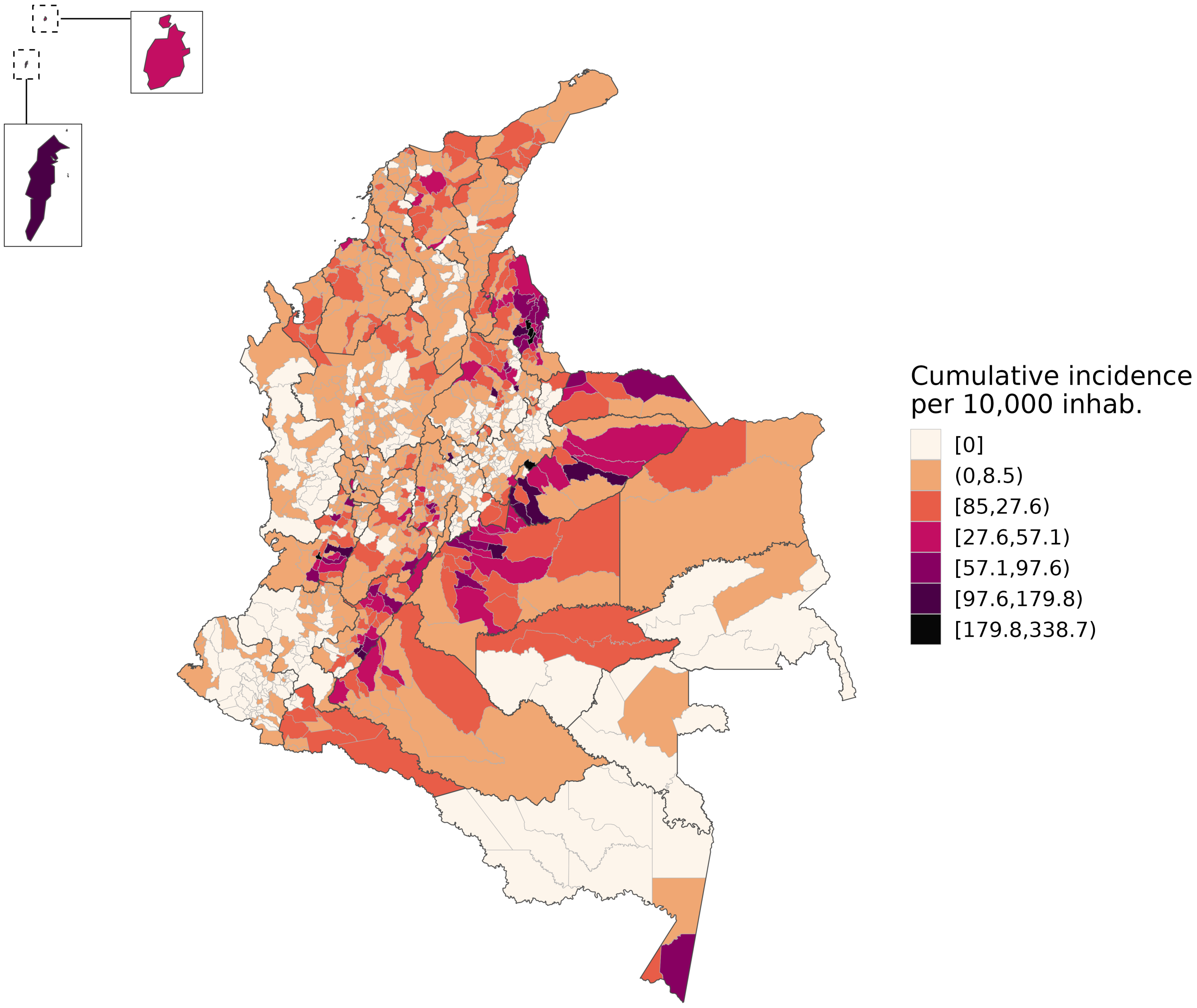


**Figure S8:** Cumulative incidence of reported Zika cases per 10,000 inhabitants by municipality of residence, epidemiological weeks 22/2015 to 39/2016, Colombia. Data source: Colombian National Public Health Surveillance System - *Sistema Nacional de Vigilancia en Salud Pública* (SIVIGILA).

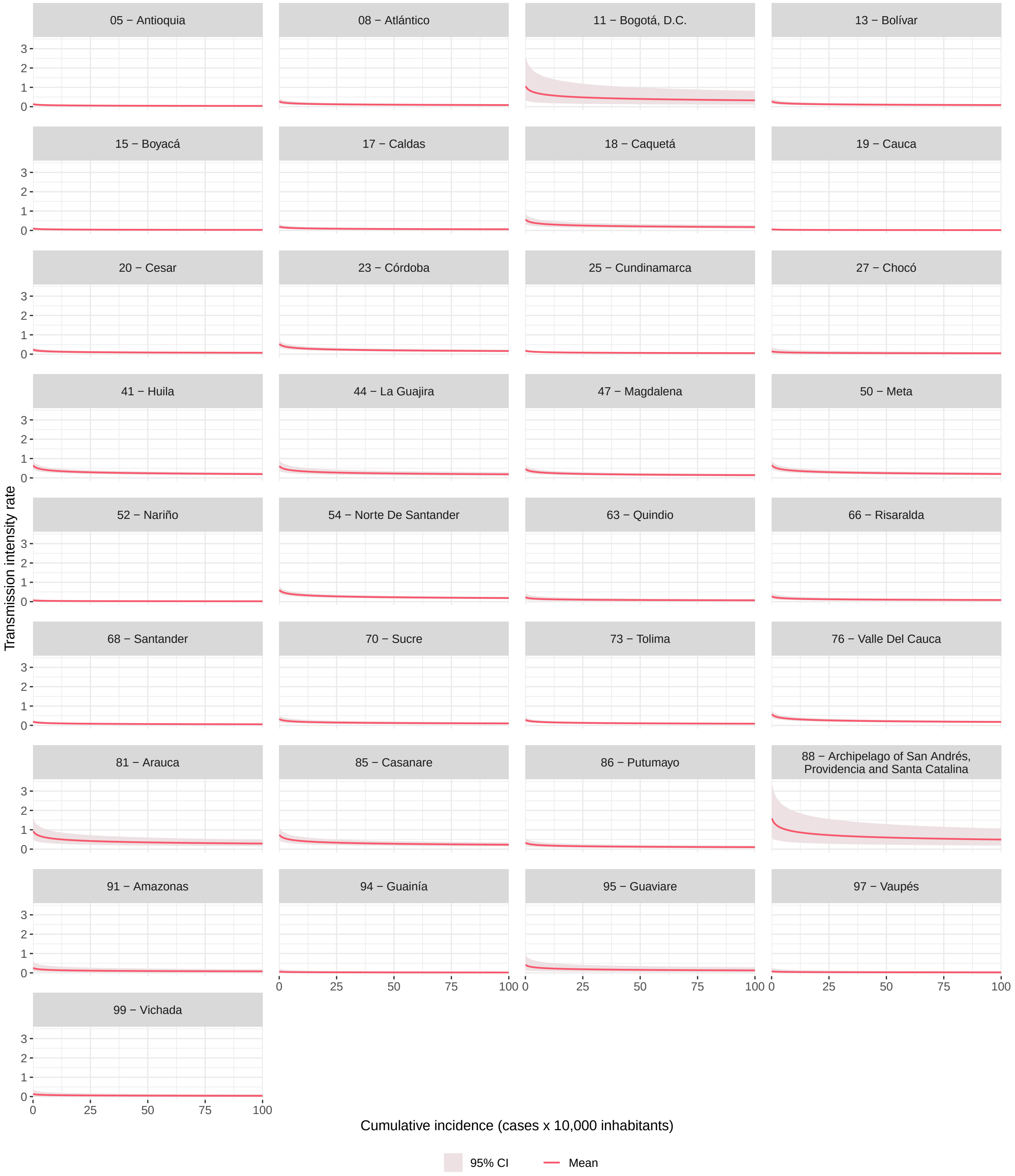


**Figure S9:** Cumulative incidence of reported Zika cases per 10,000 inhabitants (lagged by four weeks) association with the Zika transmission intensity rate by department of residence after adjusting for the department specific random effect and the average values of the other covariates in the department, epidemiological weeks 22/2015 to 39/2016, Colombia.

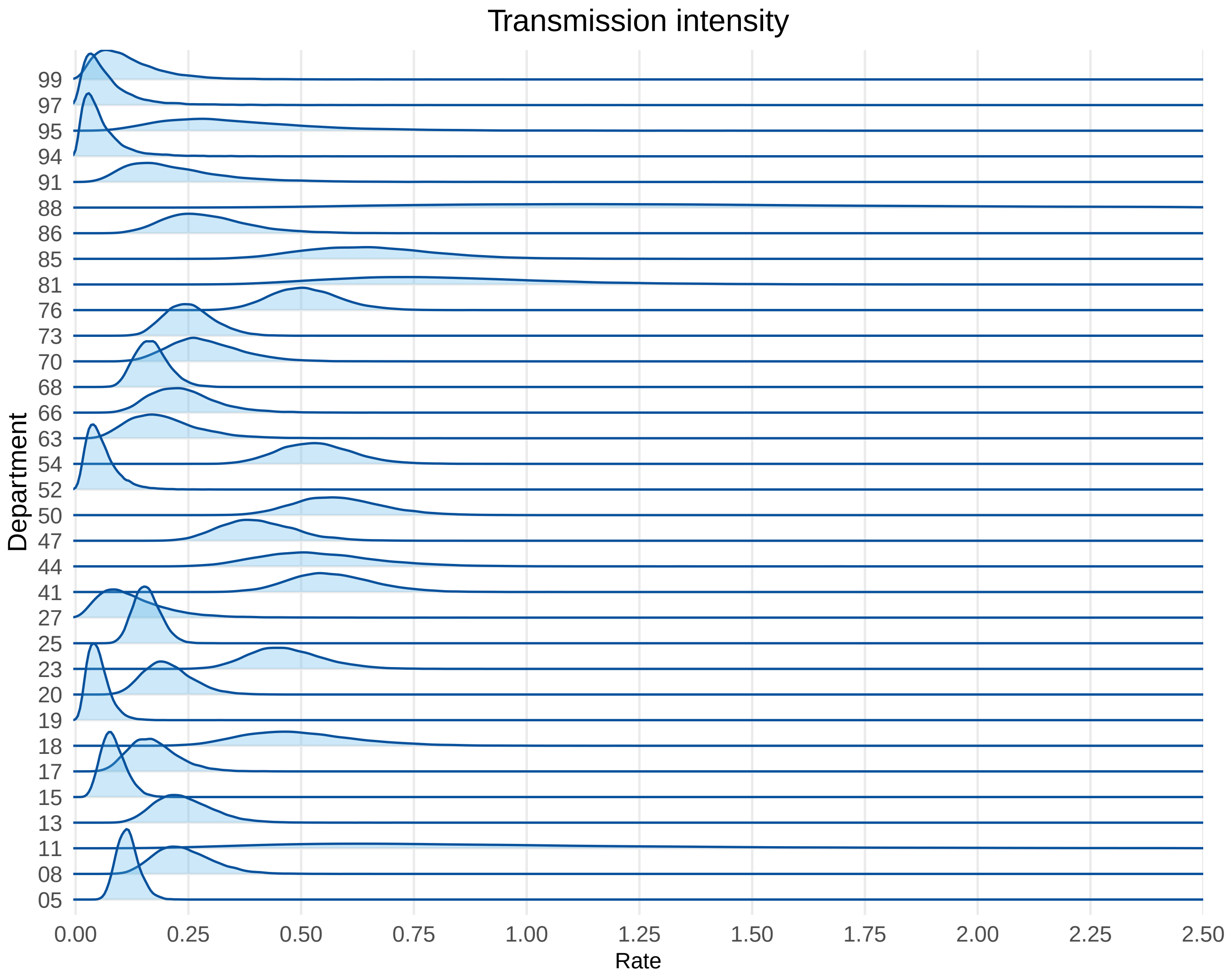


**Figure S10:** Estimated posterior distribution of the Zika transmission intensity rate by department after adjusting for the department specific random effect and the average values of the covariates in the department, epidemiological weeks 22/2015 and 39/2016, Colombia.

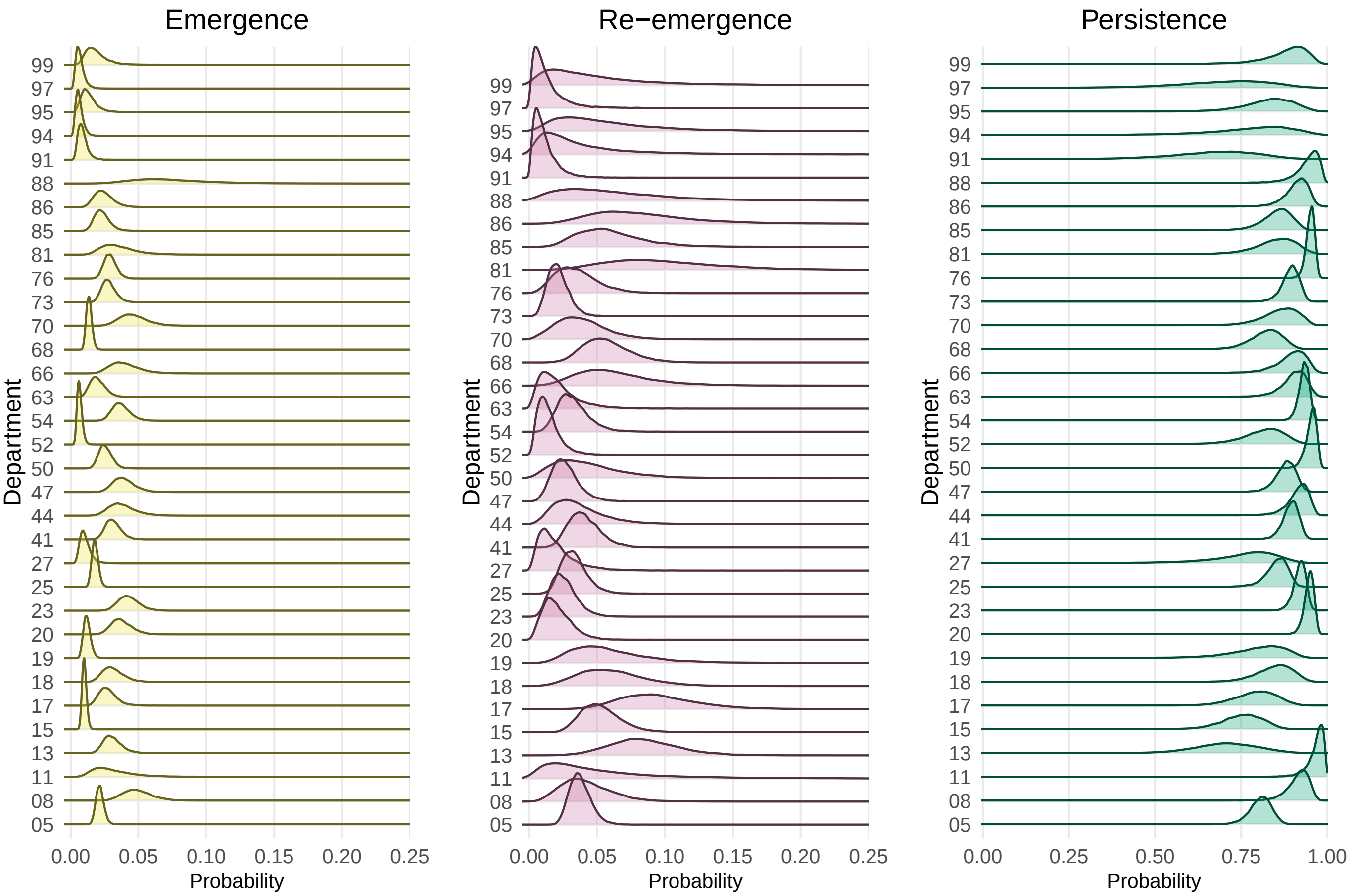


**Figure S11:** Estimated posterior distribution of the probability of emergence, re-emergence, and persistence of Zika by department after adjusting for the department specific random effect and the average values of the covariates in the department, epidemiological weeks 22/2015 and 39/2016, Colombia.

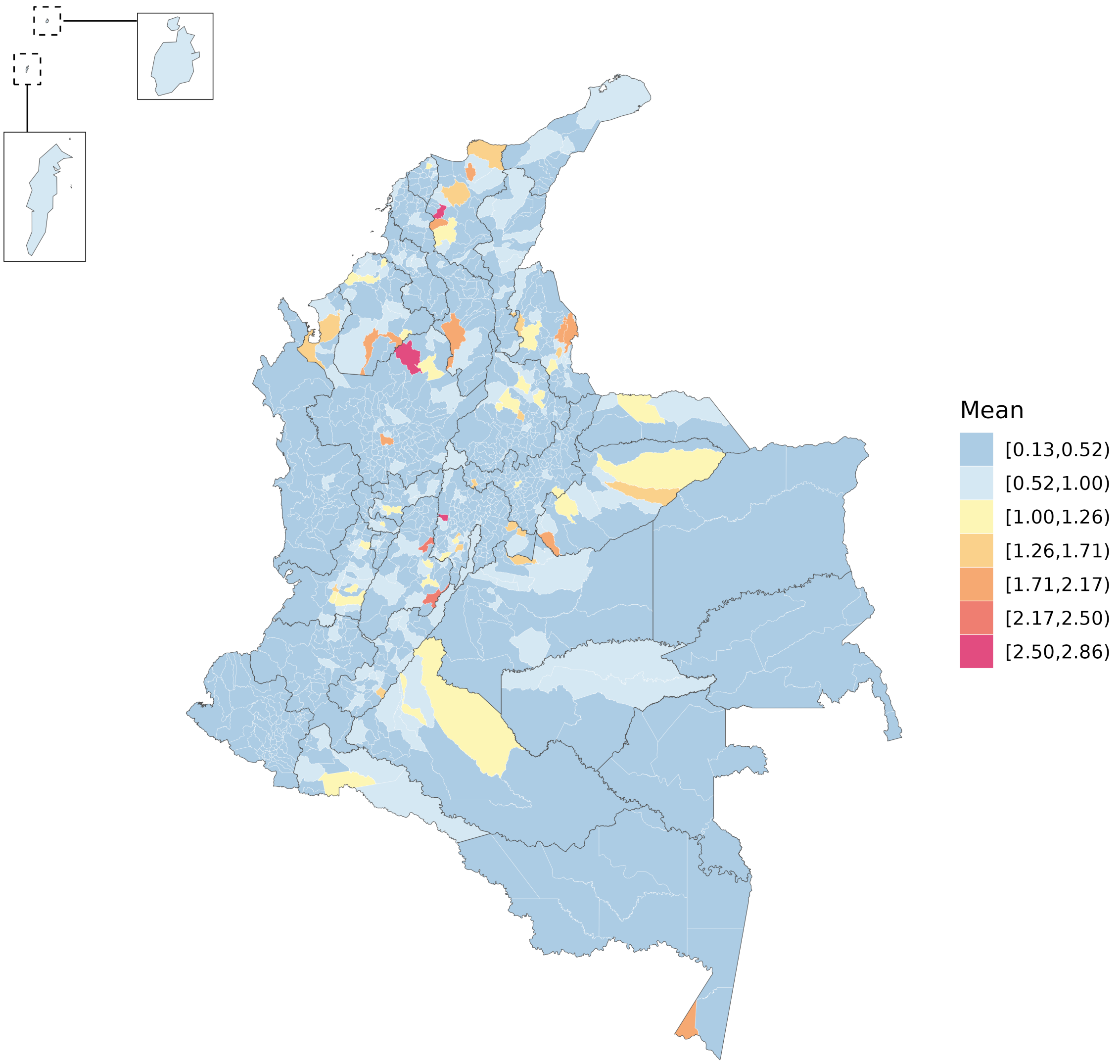


**Figure S12:** Estimated posterior mean baseline of the the expected reported cases by municipality, epidemiological weeks 22/2015 and 39/2016, Colombia.
